# A Process-Mining Model to Detect Adverse Postoperative Blood Transfusions

**DOI:** 10.1101/2022.09.21.22280177

**Authors:** Ahmet Murat Sumer, Cemil Ceylan

**Author notes:** Name and Address for Correspondence:* Ahmet Murat SUMER, *Address*: Anadolu Medical Center Hospital, Cumhuriyet Mahallesi, 2255 Sokak No.3 Gebze 41400 Kocaeli, Turkey, *Email:. Type of The Paper:* Ahmet Murat Sumer is an Industrial Engineering PhD Student in Istanbul Technical University, Turkey. This paper is created within the scope of the dissertation. The dissertation is named as “Developing a Process Mining-Based Model for Detecting Patient Safety Errors in Healthcare”. Assoc. Professor Cemil Ceylan is the advisor of this PhD dissertation. This dissertation has not been finished, yet.

## Abstract

**Importance:** Errors that threaten patient safety can cause patient harm, death, and rising health care costs. Manual process to expose adverse events (AE) increase time spend to detect them and increase costs of detection.

**Objective:** This study aims to make it easier and faster to expose AEs related with postoperative blood usages by process mining.

**Design:** These errors can be reported voluntarily by healthcare givers or exposed by Global Trigger Tool (GTT) determined by the Institute for Healthcare Improvement (IHI). With process mining ‘Transfusion of Blood or Use of Blood Products’ (C1) cases were exposed in a data set. Actual life process was discovered and GTT C1 was searched as a process pattern in the discovered process. Instead of reviewing all cases manually, only detected cases were reviewed by patient safety subject matter experts.

**Setting and Participants:** Anadolu Medical Center, Turkey was selected as the reference site for this quality improvement study. The data set includes 42,086 records, 2,870 cases and 20 activities for the period between October and December 2018.

**Main Outcomes and Measures:** With the new process mining model, data was reduced to 2,704 records, 57 cases and 16 activities. 57 cases detected by the model were analyzed by the expert group and 10 of them are defined as AEs. Rate of C1 AEs per medical record is 1.0%. The rate of C1 AEs per medical record was between 1.3% and 8.3% in other research papers.

**Conclusions and Relevance:** Instead of running the classic GTT model manually, only detected 57 patient files were analyzed. The new model 95% decreases time of experts who will review medical records to expose AEs.

## INTRODUCTION

### Patient Safety and Errors

In healthcare, ensuring patient safety is of great importance. “At least 44,000 people, possibly 98,000 people, lose their lives in hospitals each year due to preventable medical errors,” according to the results of the November 1999 study “To Err Is Human: Building a Safer Health System”^1^ published by the Institute of Medicine. This played a major role in the importance of the concept of patient safety.

According to Makary and his colleagues^2^, deaths due to medical errors in the USA are third among death causes. According to this study, 250,000 people die a year from medical errors. According to USA mortality statistics released by the Centers for Disease Control and Prevention, 611,000 people died from heart disease, 585,000 died from cancer and 150,000 died from respiratory system diseases. Deaths due to medical errors take third place among cancer and respiratory diseases in death statistics.

Patient safety problems occur while providing health care. They include transfusion errors and adverse drug events; wrong-site surgery and surgical injuries; preventable suicides; restraint-related injuries or death; hospital-acquired or other treatment-related infections; and falls, burns, pressure ulcers, and mistaken identity^1^. Prevention of these preventable medical errors is of great importance both in not harming patients and reducing cost pressure on the health system.

Healthcare givers can report patient safety incidents voluntarily. These incidents are directed to quality, patient safety, risk management functions of the institutions.

According to another study conducted in the USA, AE detection methods commonly used to track patient safety in the USA — voluntary reporting and the Agency for Healthcare Research and Quality’s (AHRQ) Patient Safety Indicators — fared very poorly compared to other methods and missed 90% of the AEs^3^.

### Global Trigger Tool (GTT)

Patient safety incident reporting made voluntarily by employees doesn’t represent the entire universe of patient safety errors. The GTT^4^, developed by IHI, is also used to expose events. Use of trigger tools is seen as an effective tool for detecting adverse events.

GTT aims to review medical records by experts to determine existence of specific trigger events. The events, triggers a case of possible AE that needs to be investigated in detail. The process of GTT is carried out with file review and expert participation.

### Postoperative Blood Transfusion

One of the trigger events identified by the IHI is C1 “Transfusion of Blood or Use of Blood Products”. According to GTT, procedures can require intra-operative transfusion of blood products for replacement of estimated blood lost, but this has become less common with “bloodless surgery.” Any transfusion of packed red blood cells or whole blood should be investigated for causation, including excessive bleeding (surgical or anticoagulant-related), unintentional trauma of a blood vessel, etc. Transfusion of many units or beyond expected blood loss within the first 24 hours of surgery, including intra-operatively and post-operatively, will likely be related to a peri-operative adverse event^4^.

A GTT application in a Turkish healthcare setting, 229 sampled patient records were collected and analyzed for a 12-months-period. 3 AEs detected and rate of C1 AEs per medical record was 1.3%.^5^

While applying the GTT in Thailand, 576 medical records reviewed. Number of blood safety AEs was 6. Rate per medical record was 1.0%.^6^

A study in China run GTT for 480 medical records only for patients over 60 years. 40 AEs detected related with C1. Rate per medical record was 8.3%. AEs in general occurred at least once in 329 (68.54%) patients. The results showed that the incidence of all-cause harm appear to be higher in Chinese elderly patients than in other groups, which the rate of patients with at least one AE was 16.60%-39.80%.^7^

While applying the GTT in German Hospitals study, 120 medical records reviewed from general surgery and neurosurgery clinics. Number of transfusion or use of blood products AEs was 9. Rate per medical record was 7.5%.^8^

### Process Mining

Studies aimed at uncovering real life from data sets have been found over the last hundred years. In the nineties, when data mining was gaining speed, processes were not given the necessary attention. The first studies on process mining are based on 2003. Process mining techniques have evolved and matured over a period. Examining time-based data sets containing process steps to determine and explore processes and checking conformity between defined processes and the actual process form the most basic Process Mining areas.^9^

Process mining, which handles data mining and process management tools together. Process mining aims to uncover, explore, control, and improve real processes from data logs that are easily accessible in today’s information systems. Topics included in process mining are automated process discovery, conformity checking, social network/organizational mining, model development, model repair, status estimation and past-based recommendations.

There are three main subjects^9^:

- ***Discovery:*** A discovery technique takes an event log and produces a model without using any apriori information. Process discovery is the most well-known process mining technique.
- ***Conformance Checking:*** An existing process model is compared with an event log of the same process. Conformance checking can be used to check if reality, as recorded in the log, conforms to the model and vice versa.
- ***Enhancement:*** The idea is to extend or improve an existing process model using information about the actual process recorded in some event log. Whereas conformance checking measures, the alignment between model and reality, this third type of process mining aims to change or extend the pre-model.

### Process Mining in the Healthcare

Process mining is used in various areas of healthcare, such as process improvement; compliance checking with guidelines, care maps and evaluation of medical processes and detecting frauds and abuses.

Application of Process Mining in Healthcare: A Case Study in a Dutch Hospital^10^, Business process analysis in healthcare environments: A methodology based on process mining^11^, Assessment of hospital processes using a process mining technique: Outpatient process analysis at a tertiary hospital^12^, Process Mining in Healthcare Evaluating and Exploiting Operational Healthcare Processes^13^, Supporting healthcare management decisions via robust clustering of event logs^14^, Systematic Methodology for Outpatient Process Analysis Based on Process Mining^15^, Monitoring and analyzing patients’ pathways by the application of Process Mining, SPC, and I-RTLS^16^ and Simulation of patient flow in multiple healthcare units using process and data mining techniques for model identification^17^ focused on improving the processes in healthcare with process mining.

Pathway Identification via Process Mining for Patients with Multiple Conditions^18^, A process mining-based investigation of AEs in care processes^19^, Declarative process mining in healthcare^20^ and Process mining-based medical program evolution^21^ are focused on defining compliance with guidelines, care maps and evaluation of medical processes.

A process-mining framework for the detection of healthcare fraud and abuse^22^ is focused on detecting frauds and abuses.

As a result, process mining has not been used to detect AEs including postoperative blood transfusion AEs.

## METHODS

### Defining the Purpose of Work

Process mining was used to increase the efficiency of trigger tools to expose undetectable patient safety errors.

The universe of Patient Safety errors include AEs voluntary reported by employees, errors detected manually using trigger tools and other events which cannot be detected.

Despite the 20-minute rule of GTT^4^, the process is time-consuming and therefore, it is not easy to reach the desired results running process completely manually causes inefficiency. GTT run on sampled medical records because of the impossibility of reviewing all manually.

The cost of finding an AE manually by medical record reviews is also important. Total cost of including patient safety experts for trigger analysis and physicians for AE detection is approximately €1,800 for a single potentially preventable AE.^23^ Such a practice is not sustainable. Therefore, samples that don’t include the entire universe, won’t be able to detect the real error universe.

With the use of “Process Mining” and “Trigger Tool” methods together, it is assumed that some of the remaining errors can be identified. Following stages explains the method.

The application model given in ***Figure 1*** shows how this can be implemented. This model consists of four stages:

**Figure 1:**
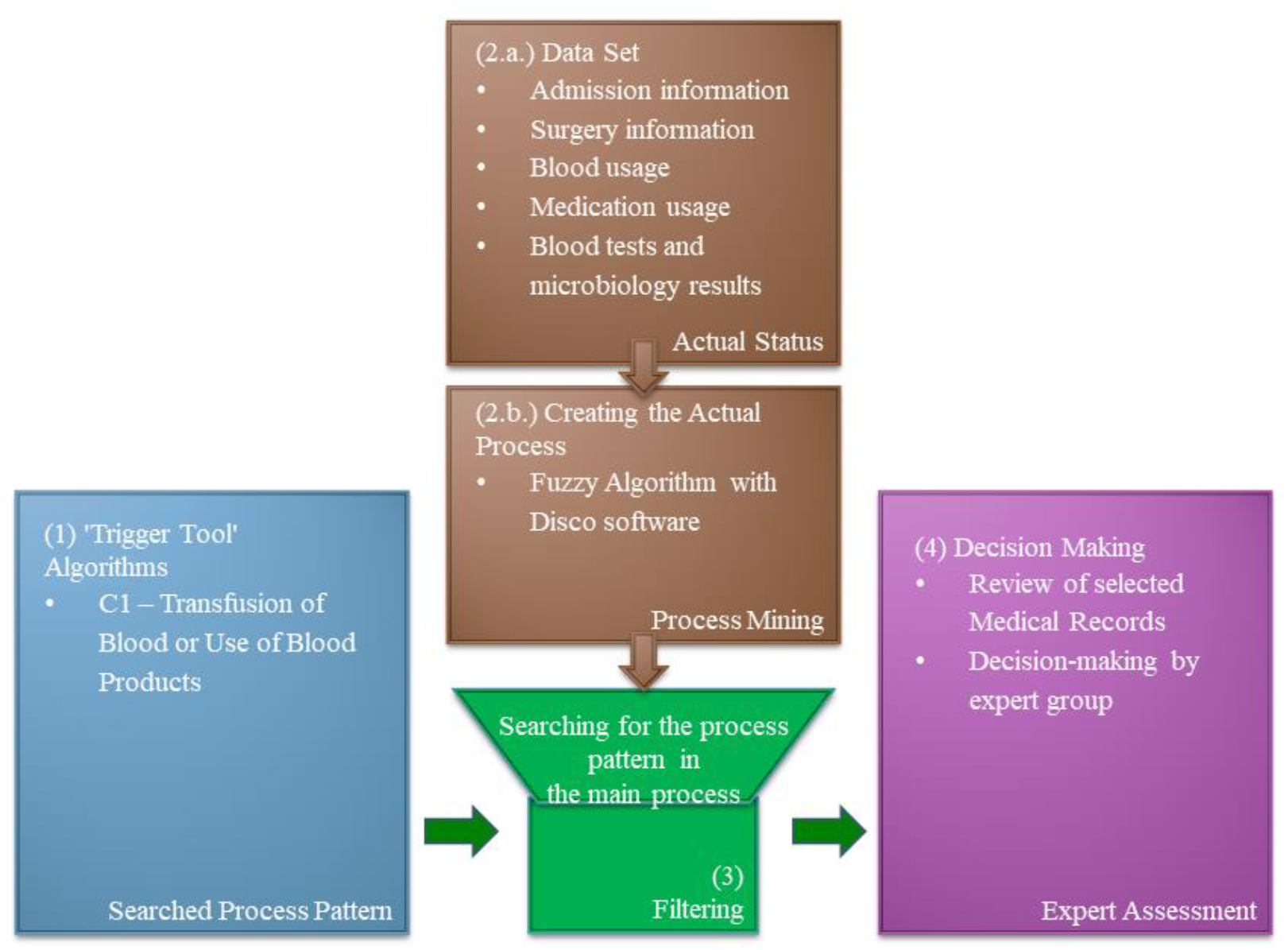
The Application Model

1. Defining the trigger tool algorithm as a process pattern
2. Creation of the data set to be examined by process mining and discovery of the actual life-process
3. Searching for process patterns in the discovered process
4. Review of the cases detected by patient safety experts

#### (1) Definition of Trigger Tool Algorithm as a Process Pattern

The trigger definitions of GTT were translated into an algorithmic structure.

- **C1 – Transfusion of Blood or Use of Blood Products**
  - Variation A
    - The patient had surgery.
    - A large amount of blood was given during the operation.
  - Variation B
    - The patient had surgery.
    - Blood transfusion was done within the first 24 hours of the surgery.

In this study, the existence of trigger tool algorithms, called process pattern, will be searched within the discovered real process.

##### (2.a) Creation of the Data Set and Collection of Data

It is essential to create the correct data set for the implementation. The data set must cover the needs of trigger pattern.

This study was carried out at Anadolu Medical Center Hospital, a healthcare institution located in Turkey, which is also affiliated by Johns Hopkins Medicine.

The diversity of institutions and Hospital Information Management Systems (HIMS) does not prevent running process mining applications. Correct configuration of the data structure is crucial.

For this purpose, HIMS was seen as the primary source. Various reports were combined into a single data set. The following list shows the types of data and sources:

- **Patient Admission and Transfer Data**
  - A report which contains patient admission and cross-departmental transfers data was created via Business Intelligence (BI).
  - According to the needed detail level on process mining, records were grouped. For instance, names of different inpatient floors are grouped as “inpatient floor”. Same was done for intensive care units.
- **Surgery Data**
  - A report which contains surgery data was created via BI.
  - Detailed surgery names were kept as additional information. Data of patients who underwent surgery or procedures were grouped as the main process step.
- **Blood Usage Data**
  - Blood and blood products administration data were reported using the HIMS.
  - The use of various blood products is grouped as “use of blood”.

Data for the period 01 October 2018 and 31 December 2018 were extracted and consolidated. Final data set includes the following fields:

- **Protocol No:** A unique and distinctive number for patients.
- **VisitID:** A unique number created by the system based on each hospitalization of the patient.
- **Source:** This shows the data source.
- **Name of the Process Item – Long:** The original name of the service provided to the patient.
- **Name of the Process Item – Summary:** The new given process item name after groupings.
- **Date:** The start date of the service.
- **Name of the Nursing Station:** Name of the nursing station where the service was given.
- **Discharge Date of the Patient:** The date of discharge.
- **Surgery Details:** Detailed name of the performed surgery.
- **Department:** The name of the medical specialty that provides the service.

##### (2.b.) Running the Process Mining Model

Various software such as PROM, Disco, etc. can be used to make process mining analyses. Disco has been preferred in this study.

When the prepared data set was uploaded to Disco, it appears that there was total 42,086 records, 2,870 cases.

Disco processes data according to its own fuzzy algorithm in default. The actual process was discovered initially. Higher number of processes and activities cause process scheme to become more complex. During this study, 20 process items were selected to reach an understandable process map. The number of paths was reduced, resulting in a simpler representation. The simplified process map (Descriptive Parameters: Activities: 54.1%, Paths: 0%) contains information about the hospital admissions, surgeries, various blood tests and blood usages.

In this process map, a process pattern formed. Process steps circled in ***Figure 2*** shows that process pattern which includes process steps in a sequence similar with the selected trigger algorithm:

**Figure 2:**
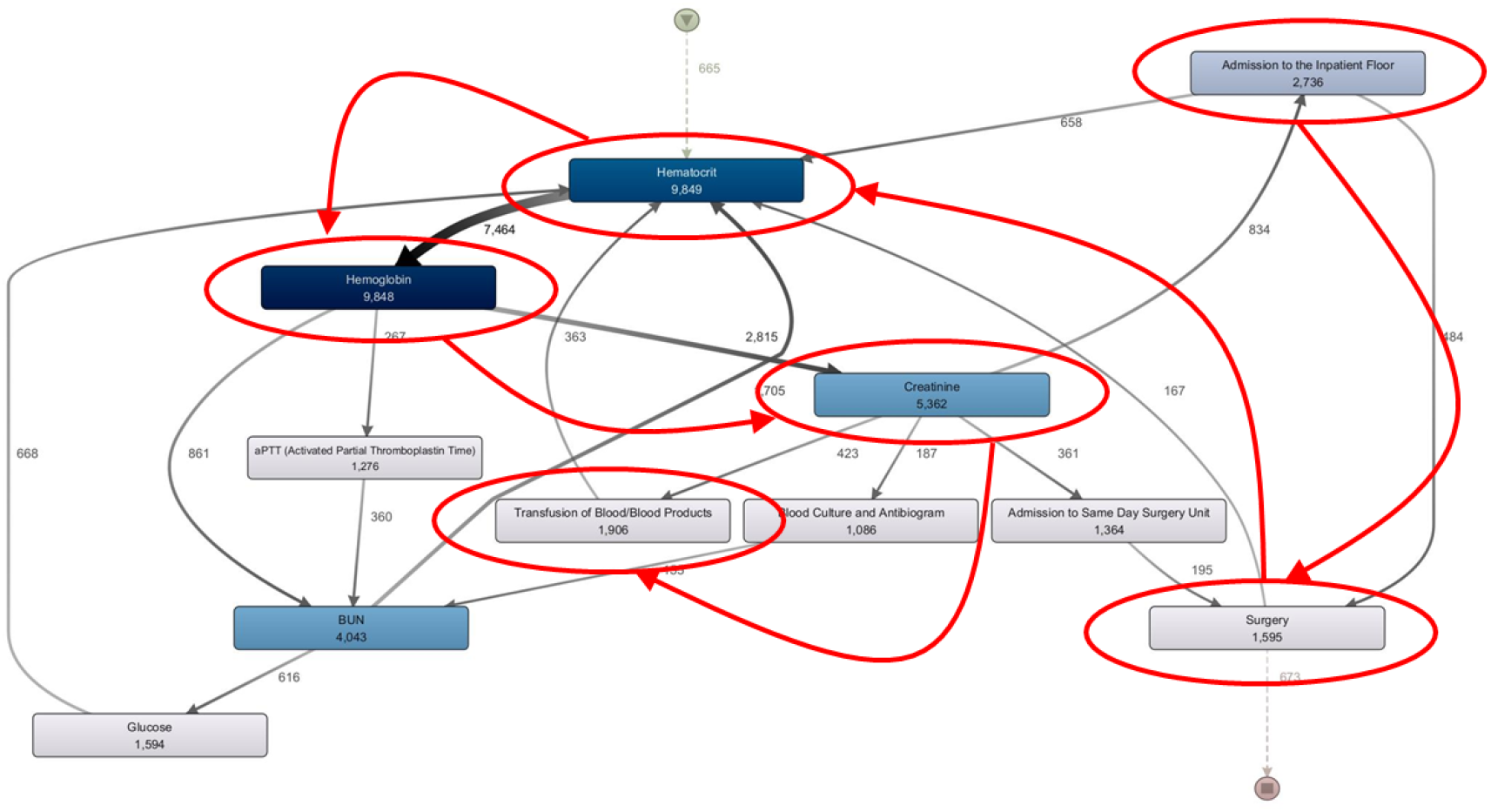
Process Pattern compatible with the trigger algorithm in the process map

1. After the patient’s hospitalization,
2. The patient had surgery,
3. Blood tests performed,
4. As a result, the patient was given blood/blood product.

According to trigger tools, the transfusion of blood or blood products within 24 hours after surgery is considered high risk in terms of patient safety. Identifying cases that fit these process pattern means revealing cases that meet the triggering conditions.

#### (3) Searching for process pattern in the main process

The above visual detection may not be able to be done visually in situations where the process and data set are complicated. Therefore, regardless of the process complexity, these process patterns can be detected with the filtering.

Rule filtering listed below must be done to define blood transfusions completed within the first 24 hours of the surgery:

- *Predecessor event:* Surgery
- *Follower event:* Blood / Blood Product Transfusion
- *The time between matching events:* Must be less than 24 hours between the two events.
- *Direct or eventually following criteria:* There may be other procedures between surgery and blood/blood product transfusion. So, the reference event must be eventually followed by the follower event.

The new process map compatible with filtering and includes cases with only defined criteria. 57 cases match this situation. There have been 182 surgeries performed and 260 blood/blood product transfusions done in these 57 cases.

#### (4) Review of the detected cases

These detected 57 cases were reviewed by patient safety subject matter experts. This review determined whether cases that met the IHI trigger criteria are an AE or not. All electronical and paper records of these patients were reviewed. After that defined 22 cases were also reviewed by a second physician reviewer.

## RESULTS

As a result of this review, the following findings on ***Table 1*** were determined. Actions to be taken regarding these have also been determined.

**Table 1:** Initial Results of Process Mining Model

| **Number of Cases** | **Findings** | **Planned Action** |
| --- | --- | --- |
| 22 | In these cases, blood use was observed within 24 hours after surgery in accordance with the trigger tool criteria. | These cases are reviewed by a second physician reviewer. |
| 21 | The duration of the surgery cannot be determined in the current HIMS. As a result, process mining model evaluated blood transfused during the surgery as postoperative blood transfusion. | Recording the start and end times of surgeries in HIMS will improve the data quality. In this case, the 24-hour period after the surgery end time can be evaluated by the model. |
| 10 | If a blood reserved on behalf of a patient is obtained from outside the institution, Turkish Red Cross (Kizilay), these reserve bloods are recorded in the Blood Bank software. In some cases, transfusion during the surgery may not be needed. However, these blood bank reservation records fell into the business intelligence software where the reporting was made, and therefore they are determined by the process mining model. | Instead of business intelligence records and Blood Bank data showing all transfusions can be used in the future analysis. |
| 4 | Stem cell collections performed in OR as a surgery. However, blood transfusion to these patients after these procedures is an expected practice. | These procedures can be excluded from data set. |

Defined 22 cases were also reviewed by a second physician reviewer. Results are shown on ***Table 2***.

**Table 2:** Results of Second Review to Define Aes

| Number of Cases | Findings | Planned Action |
| --- | --- | --- |
| 10 | The use of blood after surgery was considered worthy of investigation. In some procedures, reoperation or the presence of complications were detected. These 10 cases are defined as AEs. | It would be appropriate to review these cases by a larger group of peers at the Mortality & Morbidity Committee. This method can be used to suggest potential cases to Mortality & Morbidity Committee for peer review. |
| 10 | These were bone cancers, multi-trauma surgeries and multi-disciplinary cancer surgeries performed by Orthopedics and others. Blood usage after these complicated surgeries is generally acceptable. | - |
| 2 | These cases were catheter insertion procedures performed in OR, but they are not major surgeries. Blood transfusion was planned for these patients in the preoperative period due to the oncological problems. | These procedures can be excluded from data set. |

In the initial data set, there are a total of 42,086 records, 2,870 cases, in other words, patients, and a total of 20 transactions/activities. After running the model, this initial data set was reduced to 2,704 records, 57 cases and a total of 16 processes/activities.

According to the classic IHI trigger tool model, if this study had been done manually, 2,870 patient files would be examined individually by patient safety subject matter experts to defined TCs and by physicians for a second review to define AEs. According to 20-minute rule^4^ it must take 2,870 files x 20 minutes x 2 reviewers = 1923 hours to run GTT for all medical records. For surgical cases only, it must take 699 hours for 1,048 files. With the process mining model, reviewers must review only 57 medical records and it took 57 files x 20 minutes x 2 reviewers = 38 hours. This saves 95% of the time of the subject matter experts.

After running the process mining model and manual GTT process, 10 AEs detected related with C1 within 1,048 surgical cases. ***Table 3*** shows results and assessment. Rate of C1 AEs per medical record is 1.0%.

**Table 3:**
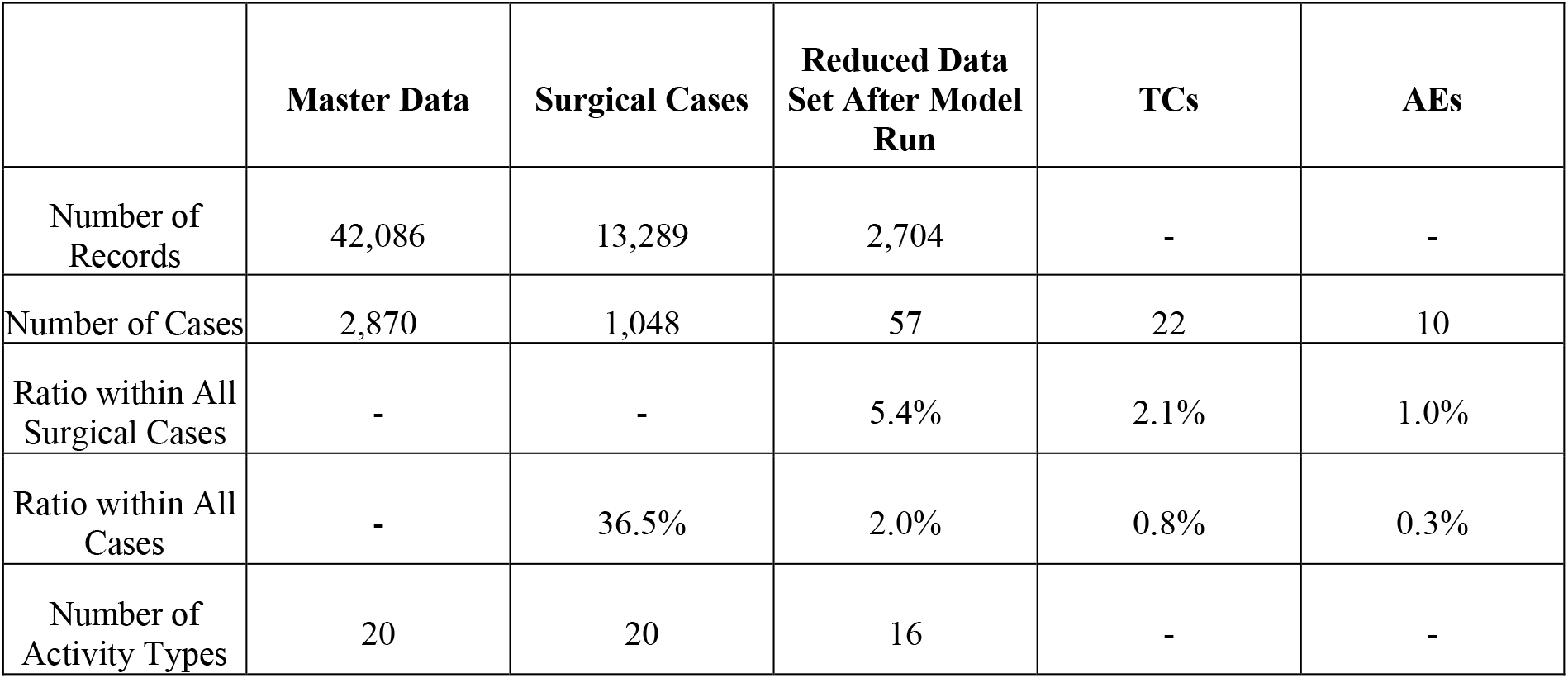
Results and Assessment

## DISCUSSION

Rate of C1 AEs per medical record is 1.0% in this study is similar with the GTT application in a Turkish healthcare setting. Rate of C1 AEs per medical record was 1.3%.^5^. However, it is below then the GTT run in China 8.3%^7^ and GTT run in German Hospitals study 7.5%.^8^ On the other hand, scope of other studies was more focused to elder patients or specific clinics then this paper. Moreover, instead of sampling, analyzing all data is a plus for this study.

Implementation of process mining in healthcare, methods for detecting patient safety incidents and trigger tool research have been discussed separately over time. However, they were not used together until this time.

Patient safety research is rapidly progressing. To achieve desired results and reduce preventable errors, sentinel events, adverse events, and near-miss events in patient safety must be accurately determined. The ability to reveal the events that cannot be detected voluntarily which constitute the iceberg’s invisible part will enable improving and preventing. As a result, patient safety risks can be mitigated.

From a macro perspective, the implementation of these studies and improving patient safety will foster the development of patient safety at the national level. Any improvement in this area will benefit both the protection of human health and the reduction of healthcare costs.

Process mining helps:

- To make of a case-based analysis,
- To search the trigger which has a process sequencing relationship,
- To analyze large data sets.

As a result, the process mining model made it possible to save 95% of the time of the subject matter experts. Instead of spending 699 hours to run a manual GTT, 38 hours will be enough. This will decrease the cost to detect AEs.

## CONCLUSION

Powering GTT with process mining model will help to reveal AEs. However, as a developing area, future studies can focus on:

- The ability of process mining to detect AEs directly with a better data set.
- Same process mining model can run for other triggers.
- Instead of detecting occurred errors, they can be prevented before by HIMS. Decision support systems and trigger algorithms should be embedded in HIMS.
- The decreased cost of detecting AEs can be calculated according to the new process mining model.
- Process mining can be used to develop new triggers.

## Data Availability

All data produced in the present study are available upon reasonable request to the authors

